# Validation of individualized flow simulations for determining the pressure gradient in patients with renal artery stenosis

**DOI:** 10.64898/2026.08.27.26361537

**Authors:** T.A. Bouwmeester, D. Collard, I.A.J. Zijlstra, E. van Hulst, A.B.G.N. Lamers, L. Vogt, B.J.H. van den Born, L. van de Velde

**Affiliations:** Department of Vascular Medicine, AmsterdamUMC, Location AMC, University of Amsterdam, Amsterdam, The Netherlands; Department of Radiology, AmsterdamUMC, Location AMC, University of Amsterdam, Amsterdam, The Netherlands; Department of Nephrology, AmsterdamUMC, Location AMC, University of Amsterdam, Amsterdam, The Netherlands; Department of Vascular Surgery, AmsterdamUMC, Location AMC, University of Amsterdam, Amsterdam, The Netherlands; Amsterdam Cardiovascular Sciences, Atherosclerosis & Aortic Diseases, Amsterdam, The Netherlands; M3i Multi-Modality Medical Imaging, TechMed Centre, University of Twente, Enschede, The Netherlands

**Author notes:** Corresponding author: Dr. L van de Velde, Department of Surgery, Amsterdam Cardiovascular Sciences, Aortic Diseases & Atherosclerosis, AmsterdamUMC, Meibergdreef 9, 1105AZ Amsterdam, The Netherlands.

**Keywords:** renal artery stenosis, renovascular hypertension, pressure gradient, computational fluid dynamics, fractional flow reserve

## Abstract

**Objectives:** To validate two computational fluid dynamics (CFD) models derived from computed tomography angiography (CTA) for estimating trans-stenotic pressure gradients, using invasive intra-arterial pressure measurements as the reference standard in patients with renal artery stenosis (RAS).

**Background:** We assessed whether non-invasive assessment of the pressure gradient using CFD could be a reliable alternative to intra-arterial measurements for identifying hemodynamically significant RAS.

**Methods:** We performed intra-arterial measurements at rest and during dopamine-induced hyperemia to assess the trans-stenotic pressure gradient in 28 patients with RAS. A pre-intervention CTA scan was used to simulate the pressure gradient with a CFD model using a strategy based on Murray’s law (CFD-Mu) and cortical volume (CFD-C). The agreement between the simulated and measured pressure gradients was assessed using intraclass correlation coefficients (ICC), Bland-Altman analysis and diagnostic agreement on the presence of a hemodynamically significant stenosis.

**Results:** In 20 patients, successful measurements and simulations were obtained. The ICC between measured pressure gradient and the CFD pressure gradient was 0.78 and 0.94 during baseline and 0.86 and 0.72 during hyperemia, for CFD-Mu and CFD-C, respectively. The sensitivity of CFD-Mu and CFD-C was 70% for both models at rest and 100% compared to the hyperemic measurements, whereas the specificity was 90% and 70% at rest and 79% and 72% during hyperemia, respectively.

**Conclusions:** The results support the use of individualized CFD simulations for hemodynamic assessment of RAS using CTA as input. The CFD models demonstrated high accuracy for the identification of a hemodynamically significant stenosis.

## Introduction

Renal artery stenosis (RAS) is a frequent cause of secondary hypertension, with prevalence escalating from <1% in mild to moderate hypertension to between 15 and 60% depending on the presence of comorbid conditions such as heart failure and renal insufficiency.[1, 2] Management focuses on optimal medical management, with endovascular revascularization reserved for high-risk clinical scenarios based on clinical and radiological grounds, including the presence of high-grade stenosis.[3] Selecting suitable candidates who may benefit from percutaneous transluminal renal angioplasty (PTRA) at an earlier stage remains challenging, in particular because different randomized controlled trials have failed to show benefit of PTRA over optimal medical therapy.[4-6] Intra-arterial pressure assessment with a vasodilator outperforms angiography alone in predicting procedural success for atherosclerotic RAS,[7-10] and some guidelines endorse them for FMD where angiographic evaluation is imprecise.[11] Yet, these invasive techniques carry risks of dissection, embolus formation or access-site hematoma, are expensive as they require a specialized team and materials, and can be technically demanding. Therefore, they have not achieved routine clinical use for this prevalent condition.[12]

Computational fluid dynamics (CFD) offers a non-invasive alternative by estimating translesional pressure gradients from a CTA using anatomic vascular reconstructions.[13, 14] If proven accurate, CFD would be an accessible method for selecting patients who likely benefit from revascularization. To date, CFD models for RAS have not integrated individualized renal blood flow rates and lack validation against invasive measurements. We hypothesized that renal flow could be estimated from the distal non-stenosed renal artery diameter, applying Murray’s law, or through a linear relation with renal cortical volume. In this study, we developed and evaluated two strategies for individualized flow assignment in CFD models of RAS and validated CFD-derived resting and hyperemic pressure predictions against in vivo measurements with a combined pressure-flow wire

## Methods

This study was designed as a prospective, single-center study. It was approved by the regional Medical Ethics Committee (METC Amsterdam UMC) and the local institutional review board. The study protocol was registered in *the Netherlands Trial Registry* (NL8408). The study was conducted in accordance with the principles of the Declaration of Helsinki, and all patients provided written informed consent.

### Study population

Clinically stable patients, aged ≥ 18 years with an indication for percutaneous transluminal renal angioplasty (PTRA) were considered for participation in the study. These included patients with suspected RAS on CT or MRA (due to atherosclerotic RAS, FMD or other cause), in combination with hypertension or a progressive decline in kidney function. Patients were eligible for inclusion if a CTA with a maximum slice thickness of 1 mm, performed within the previous 12 months, was available for use in the CFD model.

Patients were excluded if they had a history of atrial fibrillation or an estimated glomerular filtration rate (eGFR) <30 mL/min/1.73m^2^. In addition, women of childbearing age not using active birth control, and individuals deemed unable to sign informed consent due to cognitive impairment, were excluded.

Diabetes and/or hypertension were based on medical history or the use of glucose-lowering or antihypertensive medication. Blood pressure was measured using a validated 24-hour ambulatory blood pressure measurement (ABPM) device (Spacelabs 90207/90217; Spacelabs Healthcare, Snoqualmie, USA). If the 24-h ABPM measurement was of insufficient quality, an office BP measurement was used as an alternative. In that case, the mean of three seated measurements was used, after a 5-minute resting period using different validated oscillometric devices. The eGFR was calculated following the revised 2021 CKD-EPI equation.[15]

### Invasive pressure measurements

Pressure-flow measurements in the renal artery were performed as described previously.[16] Briefly, pressures were simultaneously recorded with a 6 Fr guiding sheath in the perirenal aorta and distal to the RAS lesion with a 0.014” pressure-flow wire (Combowire, Philips, Eindhoven, the Netherlands). The wire was positioned at least 5 diameters downstream of the stenosis to account for the pressure recovery phenomenon.[17] For the last three subjects a pressure-only wire (Verrata plus, Philips, Eindhoven, the Netherlands) was used due to discontinuation of the pressure-flow wire. Measurements at rest (measurement duration: 5 min) and after a vasodilating dopamine bolus of 30 µg/kg into the renal artery (measurement duration: 3 minutes) were used for this study. For RAS lesions that involved the ostium, the administration of dopamine required the pressure wire to be switched for another wire to introduce the guiding sheath or a microcatheter into the renal artery for intrarenal administration. In these cases, the pressure wire was re-introduced at the same position after dopamine administration and the sheath (or microcatheter) was retracted into the aorta. After the measurements, the lesion was treated at the discretion of the interventionalist. After the measurements, or following the intervention if performed, a drift check was performed, and the pressure wire readings were corrected accordingly. The simultaneously recorded proximal aortic pressure (Pa) and distal pressure (Pd) signals were averaged over 15 cardiac cycles. From the 15 cycle-averaged signals, the pressure gradient (Pd-Pa) was calculated.

### Computational fluid dynamics simulations

A digital twin model of a patient’s aorta and renal arteries combining anatomic and functional information was reconstructed from a CTA. Three-dimensional CFD simulations were performed to compute the flow of blood in the digital twin model in resting and hyperemic conditions. Next to an assessment of the post-stenotic disturbed flow patterns, the CFD computation directly provides the trans-stenotic pressure gradient. The following sections describe the anatomic reconstruction, the flow rate determination from CTA and the CFD computations.

From axial CTA scans with a maximum slice thickness of 1.0 mm, the sharpest reconstruction kernel available was used to reconstruct the geometry.[18] For segmentation, the level-set technique in three-dimensional (vmtkLab v1.6.1, Orobix, Bergamo, Italy) or two-dimensional centerline reconstructions was used depending on CTA quality, with manual correction and smoothing applied where necessary.[19] A three-element boundary layer was applied for the meshing of the geometries in SimVascular, with mesh convergence assessed to ensure that the pressure gradient did not change more than 1% for a doubling of mesh size.

To set the resting blood flow rate, a strategy using the renal artery cross-sectional area (CFD-Mu) and a strategy using the renal cortical volume (CFD-C) were used. For CFD-Mu, a relation based on Murray’s law[20] between the non-stenosed renal artery cross-sectional area and the flow rate was set, given by Equation 1. The constant c_1_ was fitted using the least squares method on flow velocity measurements from the present and previous cohorts.[21] The cross-sectional area A was measured distal to any post-stenotic dilatations in a healthy vessel region that was not in proximity to a bifurcation. The second strategy used the renal cortical volume measured on CTA. Using 3D Slicer, thresholding operations were used to segment the renal cortex with manual removal of major vessels. A linear relation between the renal cortical volume and the renal arterial blood flow, fitted with least squares to our cohort, was used to set the target flow rate.[22] If more than one renal artery or multiple branches were present, the flow rate was distributed over them according to Equation 1.

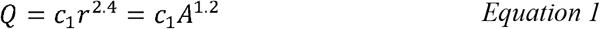

*where Q is the renal artery flow rate, r the renal artery diameter, c*_*1*_ *an empirical constant, and A the cross-sectional area*.

The hyperemic condition was simulated by reducing the peripheral vascular resistance at the outlet by a factor of 1.84, the median renal flow reserve (RFR) in our past cohorts.[21] The resulting hyperemic flow rate in the CFD simulation is dependent on the hemodynamic resistance of the stenotic lesion, with a maximum of 1.84 times the set resting flow rate for no stenotic resistance, but with lower values in case of significant stenotic resistance to flow.

The CFD model used the patient’s brachial mean blood pressure as inlet boundary condition. The renal artery outlets were set as resistance conditions, tuned to achieve the resting target flow rate within 2%, and subsequently decreased by 1.84 for hyperemic flow. Outflow in the distal aorta was assumed to be 16.67 mL/s[23]; variations to this value did not impact the RAS pressure gradient. The impact of the inlet pressure boundary condition was additionally assessed by using the invasively measured aortic pressure instead of the brachial pressure. The CFD model assumed a blood density of 1059 kg/m3, a constant dynamic viscosity set to 4.0 mPa.s, a rigid wall and steady flow, with wall motion and flow pulsatility having minimal impact on the cycle-averaged trans-stenotic pressure gradient[24]. CFD computations were performed with SimVascular version 2022.07.20 with a simulation of 1000 time steps of 0.1 ms.[25] Output data was averaged over three timepoints (0.06, 0.08 and 0.1 s) to extract the trans-stenotic pressure gradient. Figure 1 illustrates the computational workflow from CTA to the CFD model. Figure 2 presents two patient cases, atherosclerotic RAS and FMD, modelled with CFD.

**Figure 1:**
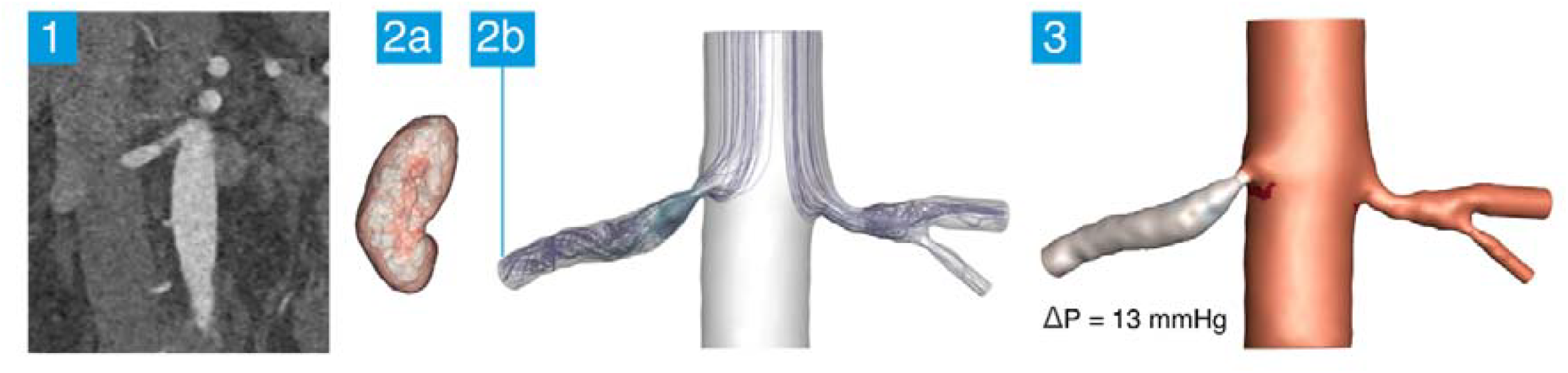
The computational fluid dynamics (CFD) model for estimating the trans-stenotic pressure gradient. (1)A computed tomography angiography.is used as input for segmenting a 3D-model of the aorta and renal arteries. (2) Two strategies using anatomical characteristics were used to set the flow rate. CFD-Cortex used the volume of the segmented renal cortex (2a), CFD-Murray used the distal renal artery cross-sectional area with Murray’s allometric scaling law (2b). The CFD model computes the motion and velocities of the blood flowing through the stenosis and provides the trans-stenotic pressure gradient, of which the resting value is shown (3).

**Figure 2:**
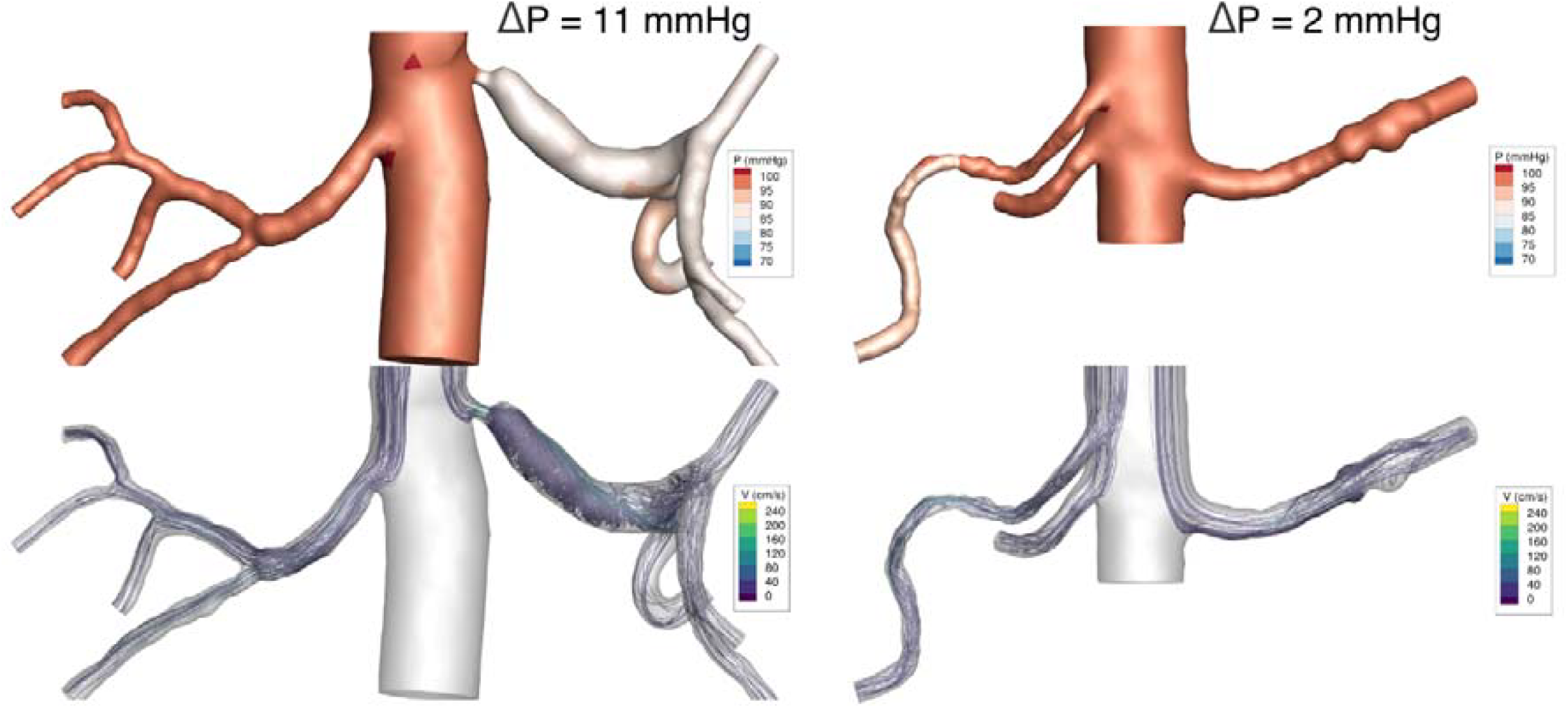
A case example of the computational fluid dynamics simulation for an atherosclerotic renal artery stenosis (left) and fibromuscular dysplasia renal artery stenosis (right) in resting conditions. P: blood pressure at the vessel wall, V: blood velocity, shown as flow streamlines in the renal artery.

### Statistical analysis

For the main analysis, we compared four modelled pressure gradients, CFD-Mu and CFD-C under both resting and hyperemic conditions, with intra-arterially measured gradients.

Resting pressure gradients from the two simulation strategies were evaluated against measured values using Bland-Altman analysis. Student’s t-test of the differences between the simulated and measured values was used to test if the mean difference was statistically different from zero. The limits of agreement (LoA) were calculated as the mean difference ± 1.96 times the standard deviation of the difference.

For each comparison we calculated the two-way intraclass correlation coefficient (ICC) for single measurements and absolute agreement, along with the Spearman correlation coefficient. ICC values were interpreted as follows: <0.50, poor agreement; 0.50–0.75 moderate; 0.75–0.90 good, and >0.90 excellent.

For comparisons between measured and modelled pressure gradients, binary classifications of significant stenosis were created based on predefined pressure gradient thresholds (≥10 mmHg at baseline and ≥20 mmHg during hyperemia).[9] We then calculated the accuracy, sensitivity, and specificity of the CFD model’s ability to correctly categorize lesions. For all analyses, p-values < 0.05 were considered significant.

## Results

A total of 30 participants were included in the study, of whom two withdrew consent prior to the intervention. Successful measurements were performed in 20 of the remaining 28 participants. A flowchart detailing inclusion and reason for measurement failure is provided in Supplemental Figure 1. Baseline characteristics of the 20 participants with successful measurements are shown in Table 1. The median age of participants was 53 years (IQR 48-60), 12 participants (60%) were men, and 16 participants were of European descent (80%) of which the majority was Dutch. All participants had a history of hypertension, with a median of 5 years since the diagnosis. Most participants (65%) used 3 or more different blood pressure lowering medications. Median eGFR was 71.5 ml/min/1.73m^2^ (IQR 40-87). Ten patients reported a history of cardiovascular disease, while one patient had a history of diabetes. One participant did not receive an ABPM of sufficient quality, therefore the office BP measurement of that participant was used in the baseline table.

**Table 1:** Baseline characteristics.

|  | Overall |
| --- | --- |
| **N** | 20 |
| Age (years, median [IQR]) | 53.0 [47.8, 59.5] |
| Sex (male, N (%)) | 12 (60.0%) |
| Ethnicity (N (%)) |  |
| European descent | 16 (80 %) |
| Other | 4 (20 %) |
| Length (cm, median [IQR]) | 175.0 [171.0, 176.5] |
| Weight (kg, median [IQR]) | 76.0 [69.0, 88.5] |
| Systolic BP (mmHg, median [IQR]) | 141.0 [129.0, 155.5] |
| Diastolic BP (mmHg, median [IQR]) | 82.5 [79.0, 92.0] |
| Time since hypertension diagnosis (years, median [IQR]) | 5.0 [1.8, 12.8] |
| Number of antihypertensives (N (%)) |  |
| 1 | 4 (20 %) |
| 2 | 3 (15 %) |
| 3+ | 13 (65 %) |
| eGFR (ml/min/1.73m, median [IQR]) | 71.5 [43.0, 88.5] |
| History of CVD (N (%)) | 10 (50.0 %) |
| Type of stenosis (N (%)) |  |
| Atherosclerotic | 8 (40 %) |
| FMD | 9 (45 %) |
| Other | 3 (15 %) |
| History of Diabetes (N (%)) | 1 (5%) |
| History of renal artery intervention (N (%)) | 6 (21.4%) |

### Hemodynamic measurements

Of the 20 participants, eight (40%) had an atherosclerotic renal artery stenosis, nine (45%) had a FMD lesion and three (15%) had a stenosis of different or undetermined etiology. The median resting pressure gradient was 9.3 (IQR 1.2-17.0) mmHg (atherosclerotic: 7.9 (IQR 2.7-13.2) mmHg; FMD/undetermined: 9.6 (IQR -0.3-18.0) mmHg). The median hyperemic pressure gradient was 7.7 (IQR 3.6-24.3) mmHg (atherosclerotic: 8.6 (IQR 2.2-17.8) mmHg; FMD/undetermined: 7.7 (IQR 4.1 – 41.0) mmHg).

### Agreement with CFD simulations

For the resting pressure gradient, the CFD-Mu modelled pressure gradient demonstrated a strong correlation with the measured pressure gradient, with Pearson’s correlation coefficient of 0.89 (p < 0.001, R^2^ = 0.80, Figure 3A) and good agreement as indicated by an ICC of 0.78 (0.52 – 0.91, p < 0.001). The cortex volume-based CFD-C simulation demonstrated a very strong correlation coefficient of 0.95 (p < 0.001, R^2^ = 0.91, Figure 3B) and excellent agreement with an ICC of 0.94 (0.87 – 0.98, p < 0.001).

**Figure 3:**
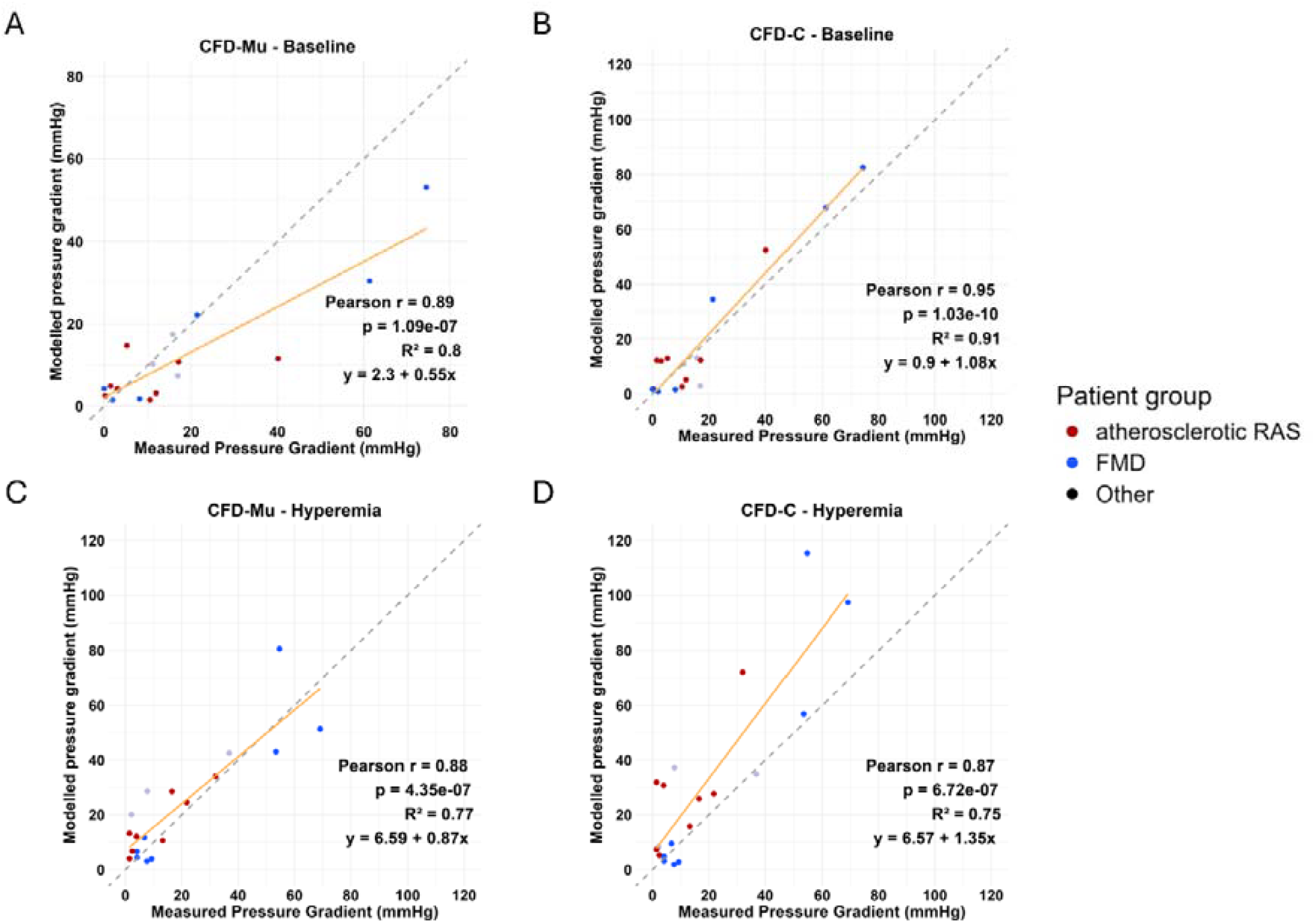
Correlation plots comparing measured and modelled pressure gradients across the renal artery stenosis. **Panel A** shows the association between the baseline measured pressure gradient and the modelled pressure gradient based on Murray’s law. **Panel B** shows the association between baseline measured pressure gradient and the modelled pressure using the cortical volume. **Panel C** shows the association between the hyperemic pressure gradient and the modelled pressure gradient based on Murray’s law. **Panel D** shows the association between the hyperemic pressure gradient and the modelled pressure gradient using the cortical volume.

During hyperemia, the CFD-Mu maintained a strong correlation with measured pressure gradients (r = 0.88, p < 0.001, R^2^ = 0.77, Figure 3C) and an ICC of 0.86 (0.68 – 0.94, p < 0.001), while CFD-C showed a similar correlation coefficient of 0.87 (p < 0.001, R^2^ = 0.75, Figure 3D), but a moderate agreement with an ICC of 0.72 (0.28 – 0.89, p = 0.002).

### Bland-Altman analysis

For CFD-Mu, Bland–Altman analysis comparing the measured and modelled pressure gradient showed a mean bias of +4.4 mmHg, indicating that the measured pressure gradient was higher than the modelled gradient (limits of agreement: –17.4 to 26.2 mmHg, p = 0.09, Figure 4A). For CFD-C, the mean bias was –2.1 mmHg, indicating that the modelled gradient exceeded the measured gradient (limits of agreement: –16.7 to 12.5 mmHg, p = 0.22, Figure 4B). During hyperemia, the mean bias was –4.2 mmHg (limits of agreement: –24.4 to 15.9 mmHg, p = 0.08, Figure 4C) for CFD-Mu, and –12.7 mmHg (limits of agreement: –47.2 to 21.8 mmHg, p = 0.004, Figure 4D) for CFD-C, showing overestimation of the hyperemic pressure gradient in both models. The use of invasively measured aortic pressure instead of estimated aortic pressure based on the brachial blood pressure from the intake visit, did not impact the accuracy of the model.

**Figure 4:**
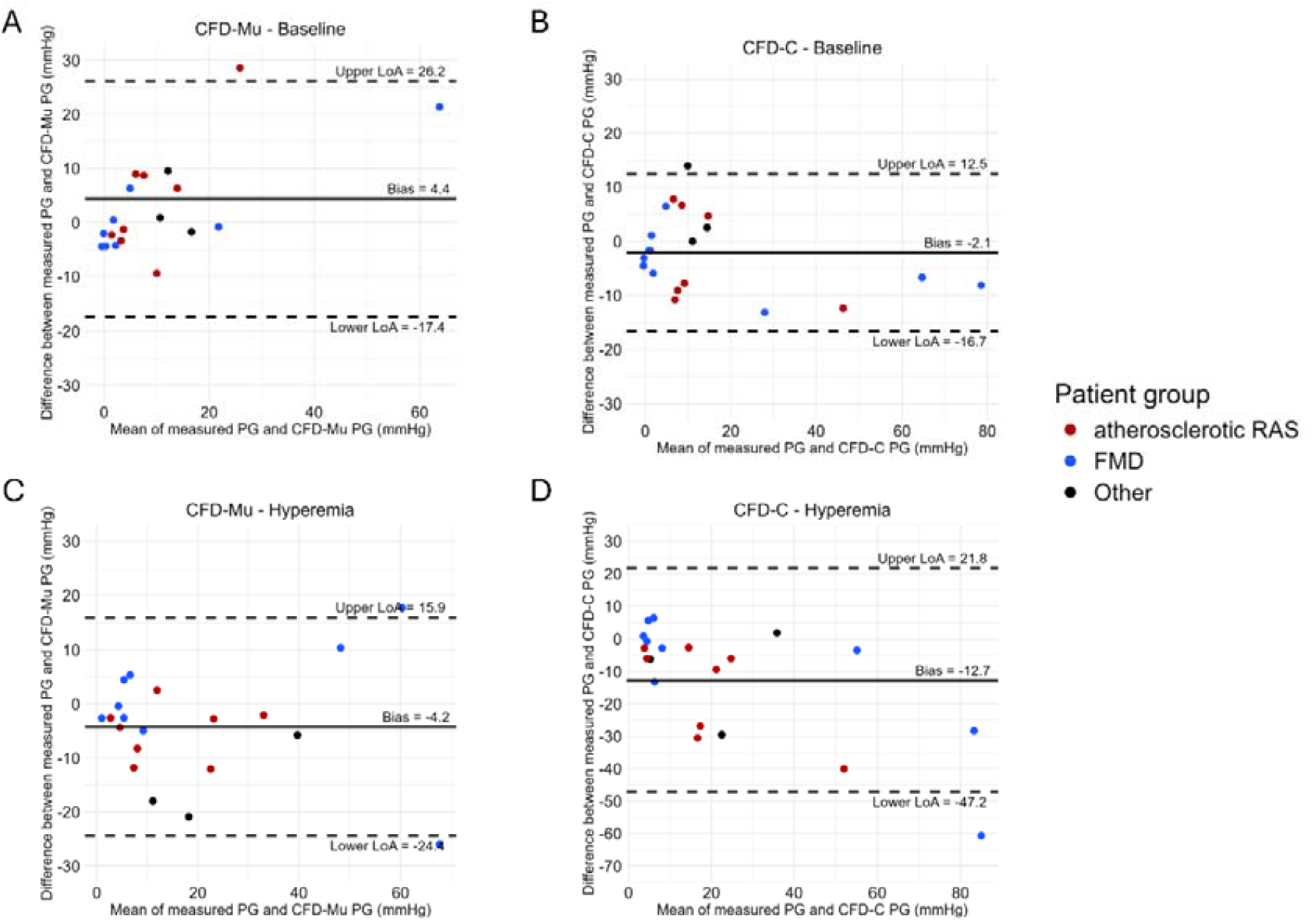
Bland–Altman plots comparing measured and modelled pressure gradients across the renal artery stenosis. **Panel A** shows the Bland–Altman plot comparing the baseline measured pressure gradient to the modelled values using Murray’s law. **Panel B** shows the Bland–Altman plot comparing the baseline measured pressure gradient to the modelled pressure gradient based on cortical volume. **Panel C** shows the Bland–Altman plot comparing the hyperemic pressure gradient to the modelled values using Murray’s law. **Panel D** shows the Bland–Altman plot comparing the hyperemic measured pressure gradient to the modelled pressure gradient based on cortical volume. PG = pressure gradient, LoA = limits of agreement, CFD = computational fluid dynamics, ARAS = atherosclerotic renal artery stenosis, FMD = fibromuscular dysplasia.

### Diagnostic accuracy

The CFD-Mu model’s classification showed a diagnostic accuracy of 80% (95% CI 56%-94%) with a sensitivity of 70% (95% CI 35%-93%) and a specificity of 90% (95% CI 55%-98%) at rest (positive predictive value (PPV) = 88% (95% CI 47%-97%), negative predictive value (NPV) = 75% (95% CI 43%-95%), while the CFD-C model’s classification showed an accuracy of 70% (46%-88%), with a sensitivity of 70% (35%-93%) and a specificity of 70%(35%-93%) (PPV = 70% (35%-93%), NPV = 70% (35%-93%)). Following hyperemia, the accuracy of the CFD-Mu model increased to 85% (62%-97%), with a sensitivity of 100% (54%-100%) and a specificity of 79% (49%-95%), (PPV = 67% (30%-93%), NPV = 100% (72%-100%), while the accuracy of the CFD-C model increased to 80% (56%-94%), with a sensitivity of 100% (54%-100%) and a specificity of 71% (42%-92%) (PPV = 60% (26%-88%), NPV = 100% (69%-100%)).

## Discussion

In this study, we demonstrate that the trans-stenotic pressure gradient in patients with RAS can be reliably approximated using a non-invasive CFD-based simulation. In our cohort, which included patients with different RAS etiologies, the CFD-estimated resting pressure gradient showed a high correlation with intra-arterial measured pressure gradients. Bland– Altman analysis demonstrated good agreement for the CFD-C model for the resting gradient, with a mean difference of -2.1 ± 7.3 mmHg, whereas the difference between CFD-Mu and the measured gradient was +4.4 ± 10.9 mmHg. The CFD error increased with higher-grade stenoses. Agreement during hyperemia remained acceptable, though the wider limits of agreement suggest increased between-method variability. However, both CFD models accurately identified hemodynamically significant stenosis under resting and hyperemic conditions. These results show that non-invasive CFD-based assessment can provide diagnostically accurate estimates of hemodynamic significance in patients with RAS.

Identifying patients with RAS who will benefit from an intervention remains challenging on clinical and radiological grounds alone. Imaging modalities such as CT angiography, magnetic resonance imaging, or ultrasound can adequately quantify anatomic stenosis severity, but have demonstrated to be poor predictors of the effectiveness of the intervention.[3] The pressure gradient of the stenosis has a stronger relation with the likelihood of a successful intervention,[9, 10, 26] with recent data confirming the wide dissociation between angiographic and pressure gradient assessment, with FFR-guidance deferring more than half of RAS lesions with a 50-90% diameter stenosis for intervention.[10]

Following hyperemia, CFD-Mu overestimated the pressure gradient by 3.6 mmHg and CFD-C by 12.7 mmHg. Limits of agreement were relatively wide, driven by cases with markedly elevated pressure gradients. Hyperemic measurements were technically more demanding and subject to greater uncertainty because the pressure wire needed to be removed from the sheath to advance the microcatheter or guiding sheath through the stenosis for dopamine delivery, after which the pressure wire was reintroduced. These additional steps may have caused pressure sensor drift or mechanical expansion of the stenotic lesion, reducing the reliability of the invasively measured hyperemic gradients and likely contributed to the lower median value of the measured pressure gradient in hyperemic compared with resting conditions.

Consequently, the extent to which the CFD model could be quantitatively validated for hyperemic pressure gradients in severe lesions was limited. Nonetheless, diagnostic accuracy for identifying a pressure gradient ≥ 20 mmHg remained high (CFD-Mu 85%, CFD-C 80%), indicating that despite potential inaccuracies in extreme cases, both models reliably identified hemodynamically significant lesions for clinical decision-making.

Overall, the present results do not clearly favor either the CFD-C or CFD-Mu model. While the CFD-C model provided a more accurate pressure gradient estimate and a higher ICC in resting conditions, the CFD-Mu model had a higher diagnostic accuracy during both resting and hyperemic conditions. Additional validation studies would be needed to support one method over the other. For now, our findings support investigating the clinical utility of both models in larger prospective atherosclerotic RAS and FMD patient cohorts with appropriate CTA and standardized follow-up of clinical outcomes.[27] Our results point to a high sensitivity of CFD and a reasonable specificity. This suggests that CFD has an excellent ability to identify responders to PTRA, but may select a small group of patients who have no significant stenosis who may not benefit from PTRA.

In the field of coronary revascularization, multiple trials have shown the added benefit of invasive coronary hemodynamic measurements compared to angiographic stenosis assessment alone.[28, 29] Consequently, CFD technology (FFR-CT) has been developed for a non-invasive assessment of coronary artery stenosis, which has seen initial clinical adoption and is supported by international guidelines.[30] For application towards renal artery stenosis, our study has substantially expanded on previous work [14] by developing individualized boundary condition strategies for CFD and demonstrating a sufficiently high agreement against invasive measurements in a group of 20 patients with RAS. Advantages of our CFD method include its lack of diagnostic burden for the patient, a standardized evaluation of hyperemic gradients and a high degree of scalability, especially if the segmentation steps are accelerated by using machine learning techniques.[31] Hemodynamic analysis using CFD to guide RAS treatment can be more readily applied in large patient studies than invasive pressure measurements, and the CFD technique also allows for retrospective analysis.

### Strengths and limitations

Strengths of this study include the use of personalized flow estimates, based on segmentation of the affected renal artery and kidney. This approach provided more accurate boundary conditions for the CFD model and yielded a resting pressure gradient with a significantly increased intraclass correlation compared to a CFD model with generalized flow input.[14] Another strength is the use of the current gold standard, namely combined intra-arterial pressure and flow measurements, for the validation of the CFD results. Several limitations should also be acknowledged. Although we corrected pressure measurements for drift at the end of each procedure, in a subset of patients 30–60 minutes elapsed between baseline measurements and the final drift check. This was necessary to preserve renal artery cannulation in technically challenging cases, but it may have introduced minor inaccuracies.

From a modeling perspective, manual segmentation of renal arteries was required, which is time consuming and currently limits clinical scalability. Development of standardized and automated segmentation tools will be essential for broader implementation. The CFD model assumed a rigid vessel wall. While the impact in atherosclerotic RAS lesions is likely minor, [24] the rigid-wall assumption may be less appropriate in multifocal FMD and warrants further evaluation. Prior studies demonstrated that pressure gradients are most sensitive to geometry and flow rate uncertainty, supporting our focus on patient-specific anatomy and flow conditions,[32] but more advanced techniques such as contrast-enhanced ultrasound or 4D-flow MRI could further refine input values at the cost of added complexity. The geometric accuracy in our model is constrained by the CTA resolution and the presence of blooming artifacts caused by calcification, though improvements are expected with the implementation of photon-counting CT.[33] Finally, our models tended to overestimate hyperemic pressure gradients, suggesting that a lower RFR value, reflecting a less pronounced reduction in microvascular resistance, may provide a more physiologic representation of renal vasodilation. Despite these limitations, our study shows that the developed CFD model has a very good ability for identifying hemodynamically significant RAS lesions and provides a foundation for future refinement and clinical translation.

## Conclusion

The developed CTA-based flow simulation of the pressure gradient in patients with RAS had a high agreement with invasively measured pressure gradients. Moreover, this non-invasive approach reliably identified hemodynamically significant lesions. CFD allows a non-invasive, CTA-based estimation of the pressure gradient and can therefore be easily applied in large patient cohorts. Future studies that investigate whether the CTA-flow simulation can predict treatment response to revascularization are indicated, to enable more effective treatment strategies in this population.

## Data Availability

All data produced in the present study are available upon reasonable request to the authors.

## Acknowledgements

This work was supported by an Innovation Grant (19OI18) from the Dutch Kidney Foundation and the authors gratefully acknowledge the Research for Health Award 2025 from the Dutch Kidney Foundation. We want to express our gratitude to all participants of our study. We want to acknowledge Jan van Haersma de With for his contributions to the study measurements.

## Data sharing statement

The datasets generated and analyzed during the current study are not publicly available due to ethical and privacy restrictions related to the use of human participant data. Data collection and handling were conducted in accordance with the approval of Amsterdam UMC METC, which does not currently provide explicit permission for unrestricted public data sharing.

Access to the data may be granted upon reasonable request, subject to approval by the Amsterdam UMC METC, and in compliance with applicable data protection regulations. Researchers who wish to access the data for replication or further analysis should submit a motivated request to the corresponding author, including a description of the intended use.

## Authors contribution

T.A.B., L.v.d.V., D.C., L.V., and B.-J.H.v.d.B. conceived and designed the research; T.A.B., L.v.d.V., D.C., IJ.A.J.Z., and A.B.G.N.L, performed experiments; L.v.d.V. constructed the CFD models; T.A.B. and L.v.d.V performed manual segmentations of renal vasculature and kidney cortex; T.A.B. and L.v.d.V. interpreted results of experiments; T.A.B. and L.v.d.V. prepared figures; T.A.B., L.v.d.V., D.C. and B.-J.H.v.d.B. drafted the manuscript; all authors edited and revised the manuscript; all authors approved the final version of manuscript.

## Supplemental material

**Supplemental Figure 1:**
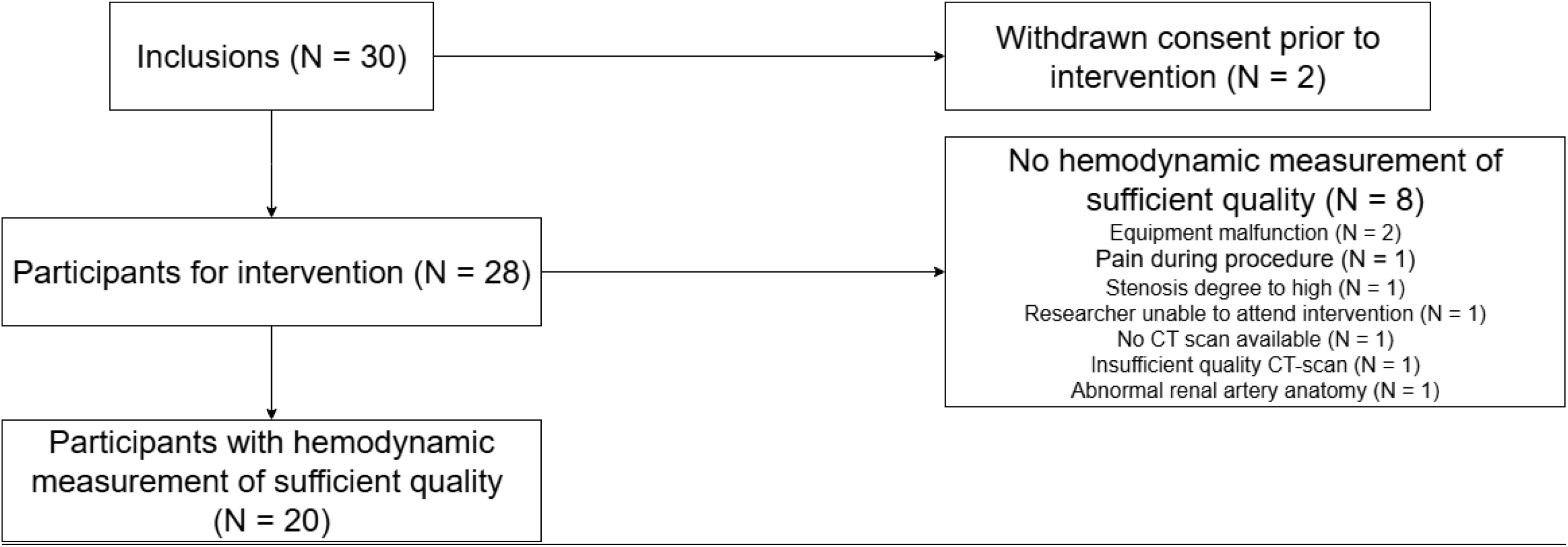
Flowchart of the inclusion of participants.

